# The disproportionate rise in COVID-19 cases among Hispanic/Latinx in disadvantaged communities of Orange County, California: A socioeconomic case-series

**DOI:** 10.1101/2020.05.04.20090878

**Authors:** Daniel S. Chow, Jennifer E. Soun, Justin Glavis-Bloom, Brent Weinberg, Peter D. Chang, Simukayi Mutasa, Edwin Monuki, Jung In Park, Xiaohui Xie, Daniela Bota, Jie Wu, Leslie Thompson, Alpesh N. Amin, Saahir Khan, Bernadette Boden-Albala

## Abstract

**Background:** Recent epidemiological evidence has demonstrated a higher rate of COVID-19 hospitalizations and deaths among minorities. This pattern of race-ethnic disparities emerging throughout the United States raises the question of what social factors may influence spread of a highly transmissible novel coronavirus. The purpose of this study is to describe race-ethnic and socioeconomic disparities associated with COVID-19 in patients in our community in Orange County, California and understand the role of individual-level factors, neighborhood-level factors, and access to care on outcomes.

**Methods:** This is a case-series of COVID-19 patients from the University of California, Irvine (UCI) across six-weeks between 3/12/2020 and 4/22/2020. Note, California’s shelter-in-place order began on 3/19/2020. Individual-level factors included race-ethnicity status were recorded. Neighborhood-level factors from census tracts included median household income, mean household size, proportion without a college degree, proportion working from home, and proportion without health insurance were also recorded.

**Results:** A total of 210-patients tested were COVID-19 positive, of which 73.3% (154/210) resided in Orange County. Hispanic/Latinx patients residing in census tracts below the median income demonstrated exponential growth (rate = 55.9%, R2 = 0.9742) during the study period. In addition, there was a significant difference for both race-ethnic (p < 0.001) and income bracket (p = 0.001) distributions prior to and after California’s shelter-in-place. In addition, the percentage of individuals residing in neighborhoods with denser households (p = 0.046), lower levels of college graduation (p < 0.001), health insurance coverage (p = 0.01), and ability to work from home (p < 0.001) significantly increased over the same timeframe.

**Conclusions and Relevance:** Our study examines the race-ethnic disparities in Orange County, CA, and highlights vulnerable populations that are at increased risk for contracting COVID-19. Our descriptive case series illustrates that we also need to consider socioeconomic factors, which ultimately set the stage for biological and social disparities.

## Introduction

By April 28, 2020, the COVID-19 pandemic has infected over 1 million individuals in the United States, with nearly 60,000 associated deaths^1^. Furthermore, COVID-19 has produced disproportionately worse outcomes among underserved minority populations. For example, recent epidemiological evidence has demonstrated a higher rate of hospital admissions and deaths among Hispanic/Latinx and African American communities^2,3^. In New York City, Hispanic/Latinx and African American patients represent 34% and 28% of fatalities despite only representing 29% and 22% of the population, respectively^4^. Understanding the critical pattern of race-ethnic disparities related to COVID-19 necessitates exploration of biological, social, and environmental factors as well as the interaction of these factors.

It is widely accepted that social determinants of health, “conditions in which people are born, grow, work, live, age, and the wider set of forces and systems,”^5^ play a critical role in the creation of disparities related to morbidity, mortality, and quality of life for most chronic, noncommunicable diseases (NCD’s) including cardiovascular disease, stroke, diabetes, asthma, and certain cancers^6^. These social determinants include poverty, wealth, educational quality, neighborhood conditions, childhood experience, social support, access, etc. Social determinants linked to NCD’s are thought to adversely impact disease outcomes via chronic exposures throughout the life course^6,7^. Unfortunately, the relatively short duration of acute infectious disease has limited investigations into the impact of social determinants, as most of this research has been conducted in chronic infections including HIV and TB^8^. However, more recent research on influenza suggested individual-level factors, neighborhood level factors, access to care, and social policies were associated with disparities in both respiratory infections and vaccine uptake^9-11^. For example, Chandrasekhar et al. observed that these factors could partially explain the variability in influenza hospitalizations with census tract data^9^. However, little is known about the social determinants of COVID-19 outcomes.

The pattern of race-ethnic disparities in COVID-19 outcomes emerging throughout the United States begs the question of what social factors may influence a highly transmissible, rapidly spreading novel coronavirus. Understanding the mechanisms associated with disparities in severe illness and death will inform the design of equitable public health strategies and preparedness plans for emerging and reemerging infectious diseases. The purpose of this study is to describe race-ethnic and socioeconomic disparities associated with COVID-19 in patients in our community in Orange County, California. Specifically, we sought to understand the role of individual-level factors, neighborhood-level factors, and access to care on COVID-19 outcomes. Further, we sought to understand the role of social distancing as an equitable public health policy that may not be available to those who cannot work from home.

## Methodology

### Patient selection

This descriptive case series included patients with confirmed COVID-19 infections who presented to the University of California Irvine Medical Center (UCI), the only academic teaching hospital (411 beds) located in Orange County, California, from March 12, 2020, to April 22, 2020. Study Day 1 was March 12, 2020, which corresponds to the date of presentation of the first confirmed COVID-19 case at UCI. COVID-19 status was diagnosed by nucleic acid detection assays for all patients. Retrospective collection of patient data was approved by UCI’s Institutional Review Board.

### Individual-level factors

We obtained demographic and socioeconomic information through manual retrospective chart review of all COVID-19 cases. Race-ethnicity data is commonly collected at UCI through patient/proxy self-report. Additional individual-level factors included the patient’s marital status, address, and occupation status. Patient home addresses were used to determine the census tract and whether the patients lived in a house or apartment.

### Neighborhood-level factors

We obtained ecological-level information through address linkage at the level of the census tract (the smallest census geographic unit in the United States) to the 2019 Federal Financial Institutions Examination Council (FFIEC) census reports, which contains 583 census tracts in Orange County^12,13^. Census tracts are uniquely numbered geographic areas that average 4,000 inhabitants from 1,600 housing units and represent relatively homogenous population characteristics^14^. As UCI is a tertiary referral center, patients geocoded to a census tract other than Orange County were excluded.

Census variables included: (1) median household income, (2) average household size, (3) percent of inhabitants 25-years and over with a college degree, (4) percent of working inhabitants 16-years and over that work from home, and (5) percent of inhabitants without health insurance. We ecologically classified individuals living in areas where the household income fell between 80% and 120% of the Orange County Median Family Income ($89,398 for 2019) as *middle-income* status^12^. We ecologically classified individuals living in areas where the household income fell below 80% of Median Family Income as *lower-income* and individuals living in areas where the household income was above 120% of Median Family income as *upper-income*. We ecologically classified the proportion of inhabitants who work from home as the ability to shelter in place.

### Statistical Analysis

Descriptive statistics were generated for demographic variables of all patients, including age, gender, and race-ethnic status. Growth patterns between weeks 1 and 6 for income brackets (*Lower, Middle, and Upper*) and race-ethnic groups (*White, Asian, and Hispanic/Latinx*) were studied by using least-squares regression analysis. We fully acknowledge that both Latinx and Black populations have been disproportionately affected by COVID-19; however, Black individuals account for 2.1% of the Orange County population^12^ and we do not have a large enough representation for statistical analysis of this group. Three models were tested to fit each time series: linear, exponential, and logistic growth curves. The slope-intercept equation, *y = mx + b*, where *y* is the number of new COVID-19 cases, *m* is the slope, and *b* is the y-intercept, was used for linear regression. The standard exponential growth equation, *a* = *Ne^rt^*, where *a* is the number of new COVID-19 cases, *N* is a constant, *r* is the growth rate, and *t* is time after the start of our collection period was used for exponential regression. A standard logistic growth equation, *a* = *a*_max_/(1 + *Be^−rt^*), where *a*_max_ is the asymptotic maximum number of articles, and *B* and *r* are constants that affect the contour of the growth curve, was used for logistic regression. A *P* value of less than or equal to 0.05 was considered to indicate significance. The Akaike information criterion was used as a measure of the goodness of fit of the three models, with the best fit determined by lowest value^15^.

To assess difference between income and race-ethnic status with respect to California’s statewide stay-at-home order (which began on 3/19/2020), proportions of cases from 2 week blocks from the pre-shelter period (Weeks 1-2, 3/12/2020 - 3/25/2020) and post-shelter period (Weeks 4-5, 4/2/2020 – 4/15/2020), were compared using Pearson’s chi-square.

Pearson’s chi-square was also used to test for significant differences in race-ethnic breakdown between county demographics, all positive COVID-19 patients, and critical COVID-19 patients, defined as need for ICU-level care or mechanical ventilation or death. Number of cases of COVID-19 per 100,000 population and proportion with critical disease were computed for each race-ethnic group with corresponding 95% confidence intervals. A *p*-value of 0.05 was considered significant for all analyses.

## Results

### Subjects

A total of 1,940 COVID-19 nucleic acid detection tests were conducted over the study period, of which 10.8% (210/1,940) were positive. Of positive cases, 73.3% (154/210) had an identifiable medical record and met inclusion criteria. This included 80 males and 74 females with an average age of 44.7 years (range, 13-88 years) (**eTable 1**).

**eTable 1.** Study population demographic information. Median household income of corresponding census tracts was available for 150 patients. Individuals living in areas where the household income fell between 80% and 120% of the Orange County Median Family Income ($89,398 for 2019) as *middle-income* status^12^. Individuals living in areas where the household income fell below 80% of Median Family Income as *lower-income* and individuals living in areas where the household income was above 120% of Median Family income as *upper-income*.

| Demographic information | No. (%) |
| --- | --- |
| **Total No.** | 154 |
| **Age, Mean [range], y** | 44.7 [13-18] |
| **Sex** |  |
| **Female** | 74 (48.1) |
| **Male** | 80 (51.9) |
| **Race-Ethnicity** |  |
| **Hispanic/Latinx** | 62 (40.3) |
| **White** | 37 (24.0) |
| **Asian** | 33 (21.4) |
| **Black** | 4 (2.6) |
| **Other / Unknown** | 18 (11.7) |
| **Household income of census tracts** |  |
| **No.** | 150 |
| **Median [range], \$** | 80,190 [25,705 – 214,323] |
| **Upper income** | 68 (45.3) |
| **Middle income** | 38 (25.3) |
| **Lower income** | 44 (29.3) |

### Individual-level Factors

The race-ethnic distribution of our patients was 40.3% (62/154) Hispanic/Latinx, 24.0% (37/154) White, 21.4% (33/154) Asian, 2.6% (4/154) Black, and 11.7% (18/154) Other/Unknown, and changes in race-ethnic distribution were monitored over the six-week study period (**Figure 1A**). During this period, the number of Hispanic/Latinx COVID-19 cases grew linearly (R^2^ = 0.997) starting after the first case in Week 2 with 5.9 new patients per week (*p* < 0.001). No significant growth pattern was observed for either White or Asian persons (*p* = 0.53 and *p* = 0.43, respectively). There was a significant difference observed in race-ethnic distribution relative to the time of shelter (Weeks 1-2 vs. Weeks 4-5) with *p* < 0.001 (**Figure 1B**).

**Figure 1.**
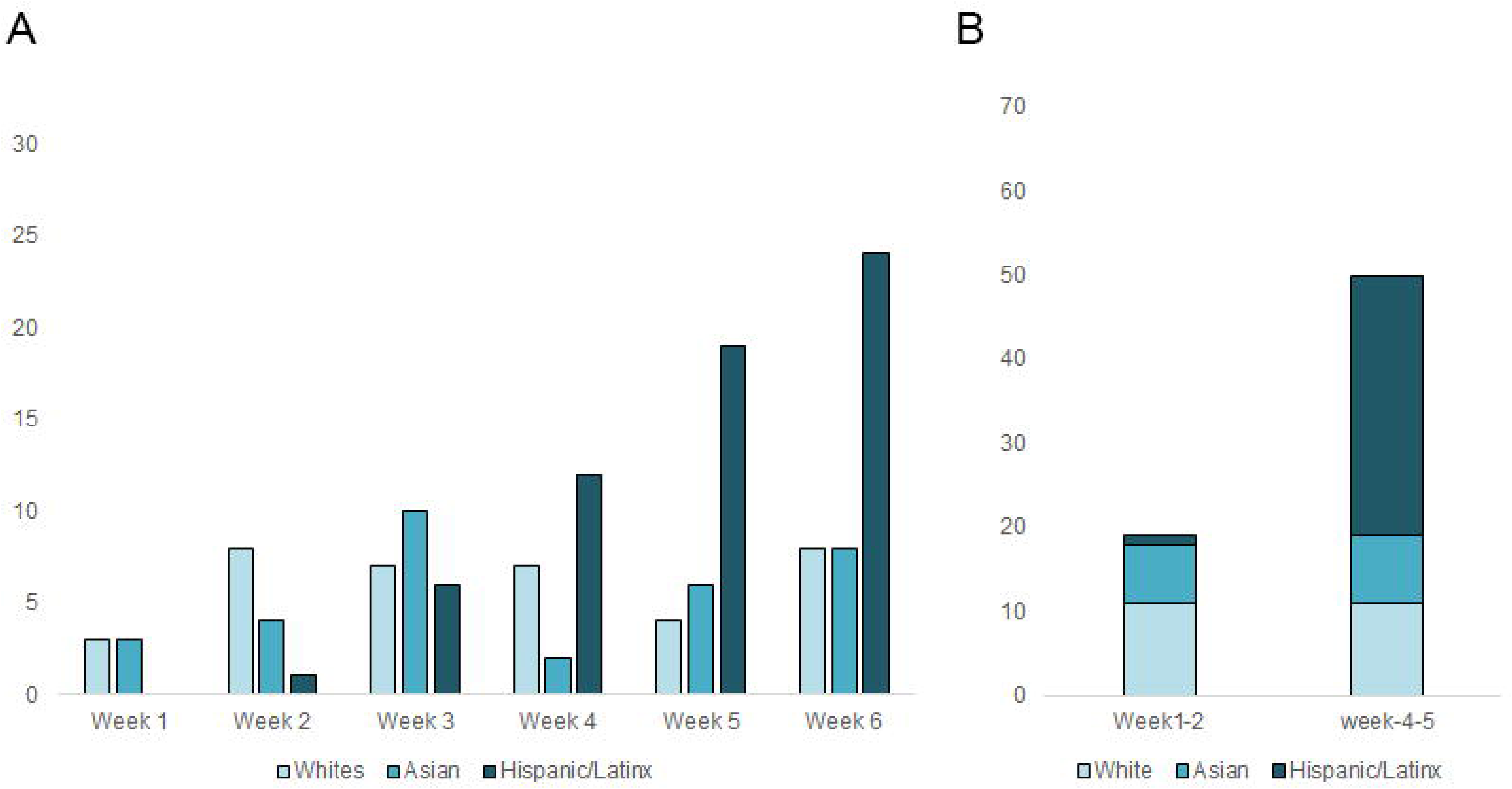
COVID-19 race-ethnic trends. Bar charts demonstrating trends for White, Asian, and Hispanic/Latinx patients from Week 1 (3/12/2020 – 3/17/2020) through Week 6 (4/16/2020-4/22/2020) demonstrating a linear increase in Hispanic/Latinx patients with other race-ethnic groups demonstrating no significant growth trend (**A**). This likely accounts for a significant difference in race-ethnic distributions relative to the time of shelter (Weeks 1-2 vs. Weeks 4-5) (**B**).

### Neighborhood-level factors

The median household income for recorded census tracts was $80,190 (range $25,705 to $214,323), which included 45.3% (68/150) Lower, 29.3% (44/150) Middle, and 25.3% (38/150) Upper income brackets over the six-week study period (**Figure 2A**). Across the study period, patients residing in low income tracts demonstrated an exponential growth rate (r = 64.2%, R^2^ = 0.958) (*p* = 0.001). Patients residing in middle-income tracts demonstrated a linear growth rate (1.9 patients per weak, *p* = 0.02). No significant growth rate was observed for patients residing in high-income tracts (*p* = 0.40). There was a significant difference observed in the income bracket distribution of census tracts relative to the time of shelter (Weeks 1-2 vs. Weeks 4-5) with *p* = 0.04 (**Figure 2B**).

**Figure 2.**
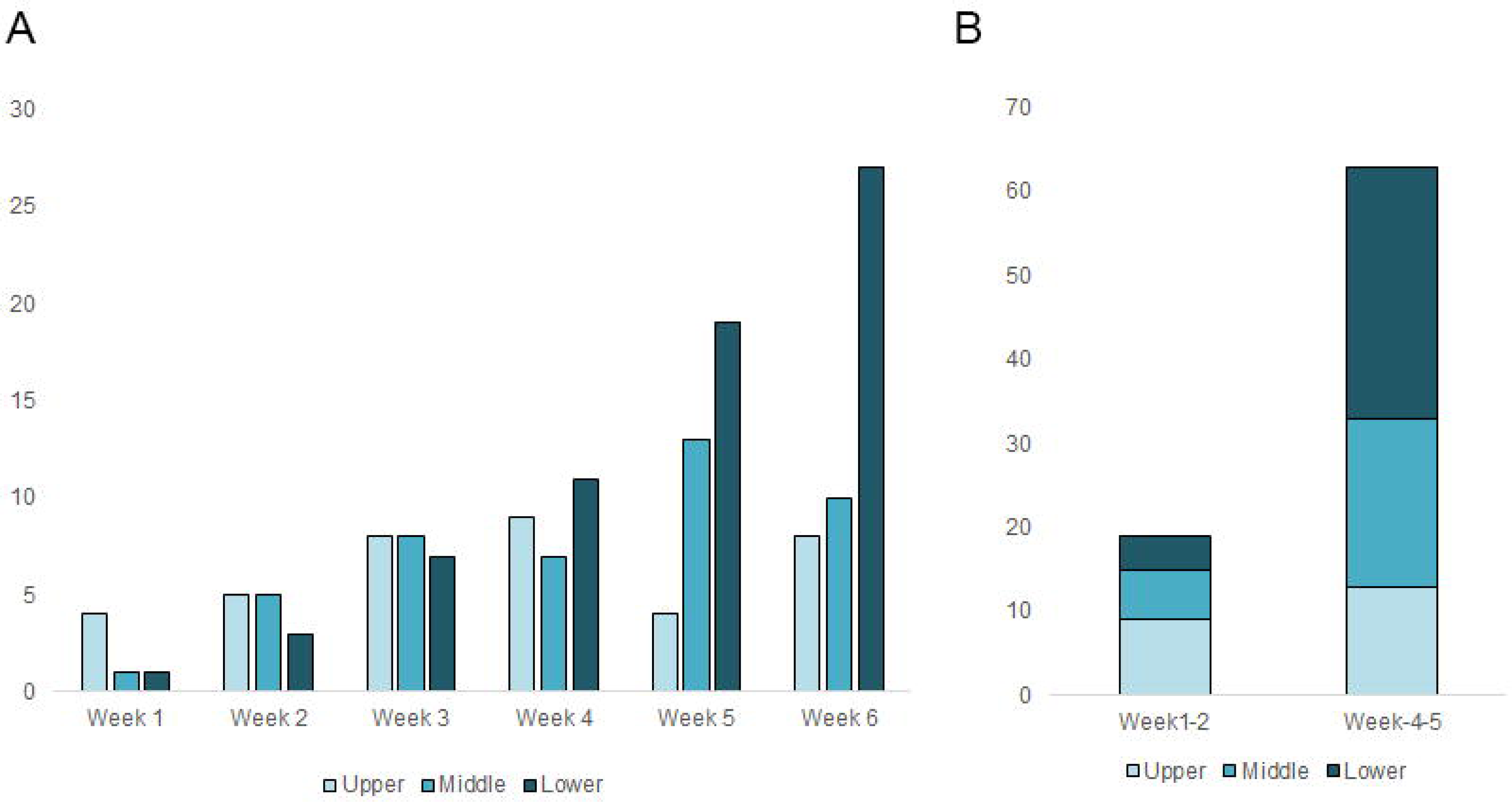
COVID-19 census tract trends. Bar charts demonstrating trends for Upper, Middle, and Lower income neighborhoods of patients from Week 1 (3/12/2020 – 3/17/2020) through Week 6 (4/16/2020-4/22/2020) demonstrating an exponential increase for lower income groups, linear increase for middle income groups, and no significant trend for upper income groups (**A**). This likely accounts for a significant difference in income bracket distributions relative to the time of shelter (Weeks 1-2 vs. Weeks 4-5) (**B**).

We divided patients into groups residing in tracts above and below the median income and examined the race-ethnic distribution of COVID-19 cases over the study period. Only Hispanic/Latinx COVID-19 cases residing in tracts below the median income demonstrated exponential growth (r = 55.9%, R^2^ = 0.9742) (**Figure 3**). We observed no significant growth rate in new COVID-19 cases amongst Hispanic/Latinx patients who lived in tracts above the Orange County median income (>$85,398) or amongst White or Asian patients in either income group.

**Figure 3.**
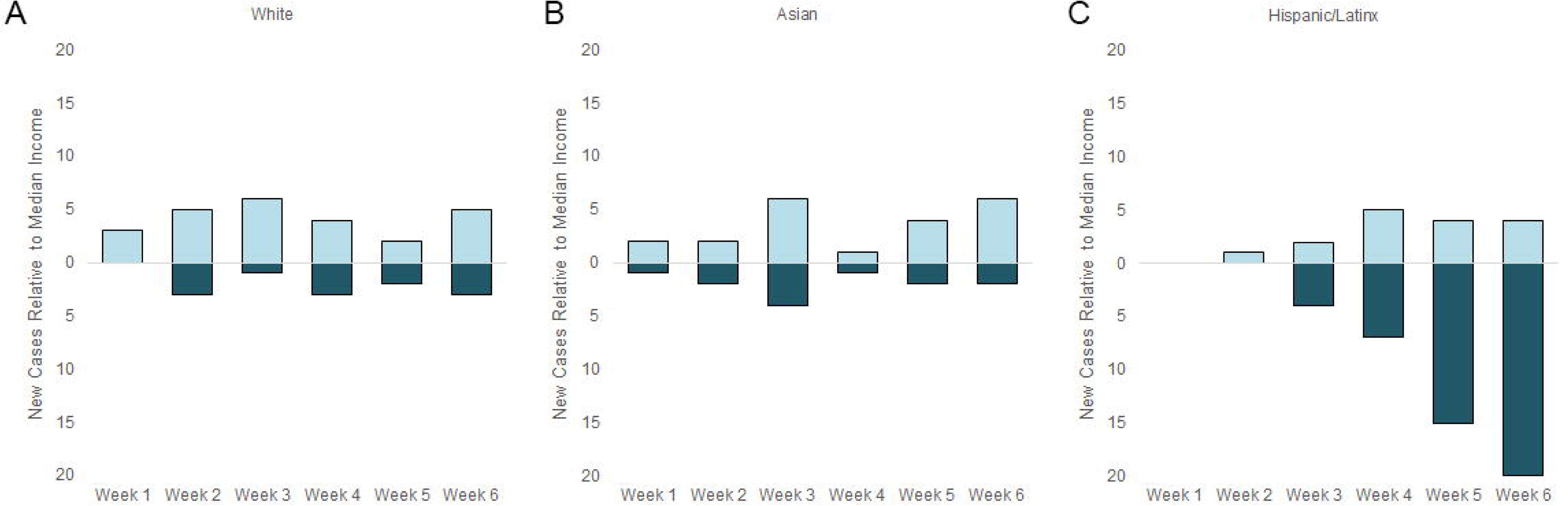
COVID-19 race-ethnic trends by census tracts. White (**A**), Asian (**B**), and Hispanic/Latinx (**C**) growth trends residing in tracts above and below the median income between Week 1 (3/12/2020 – 3/17/2020) through Week 6 (4/16/2020-4/22/2020). The only group to demonstrate a significant growth pattern was Hispanic/Latinx patients living in census tracts below the median income.

Over the study period, the mean household size assigned by census tracts rose linearly from 2.52 to 4.01 (0.28 increase per week, R^2^ = 0.903, *p* = 0.046) (**Figure 4A**). With regards to education, the mean percentage of census tract inhabitants without a college degree rose linearly from 45.2% to 80.0% (7.2% increase per week, R^2^ = 0.939, *p* < 0.001) (**Figure 4B**). The mean proportion of inhabitants working from home in the representative census tracts declined linearly from 6.3% to 3.3% (−0.6% increase per week, R^2^ = 0.989, *p* < 0.001) (**Figure 4C**). Lastly, the mean distribution of inhabitants who have no health insurance in the representative census tracts rose grew exponentially from 7.0% to 13.8% (r = 12.2%, R^2^ = 0.867) (*p* = 0.01) (**Figure 4D**).

**Figure 4.**
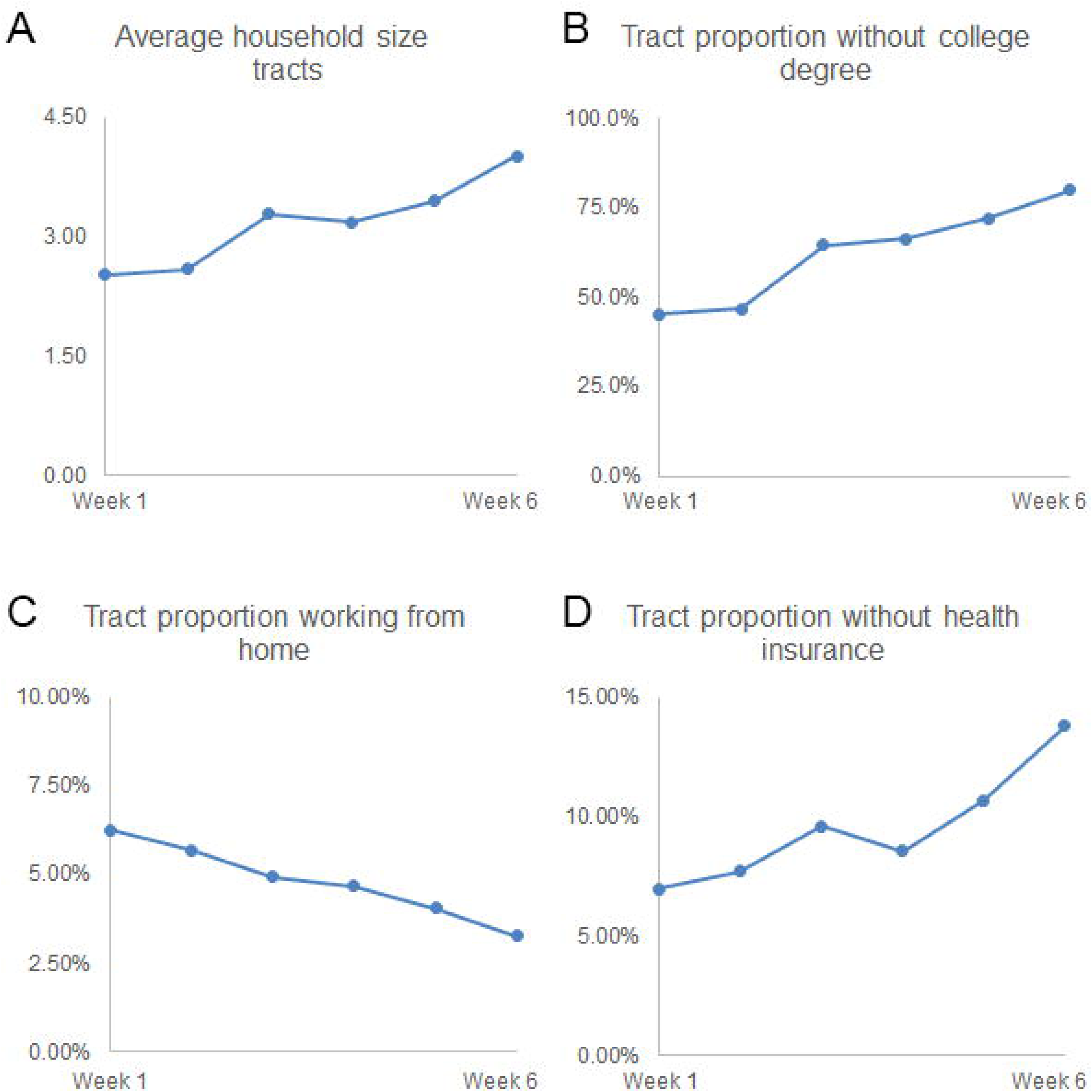
COVID-19 socioeconomic trends. Scatterplots demonstrate a linear increase in average household size (**A**), linear increase in proportion of inhabitants without a college degree (**B**), linear decrease in proportion of inhabitants working from home (**C**), and exponential increase proportion of inhabitants without health insurance (**D**).

### County Disparity

We observed significant disparities in race-ethnic distribution among COVID-19 cases with non-critical and critical disease when compared to county demographics (**Figure 5**). Compared to Orange Country’s race-ethnic distribution (40.1% White, 21.4% Asian, and 34.2% Hispanic/Latinx)^12^, the non-critical cases were comprised of 24.4% White, 20.7% Asian, and 39.3% Hispanic/Latinx and the critical cases were comprised of 21.1% White, 26.3% Asian, and 47.4% Hispanic/Latinx. In addition, the COVID-19 cases per 100,000 people were also disparate among race-ethnic groups, with 2.9 (95% CI: 0.71 to 6.8) White cases, 3.0 (95% CI: 1.4 to 6.1) Asian cases, and 9.4 (95% CI: 7.2 to 12.6) Hispanic/Latinx cases per 100,000 people of each group.

**Figure 5.**
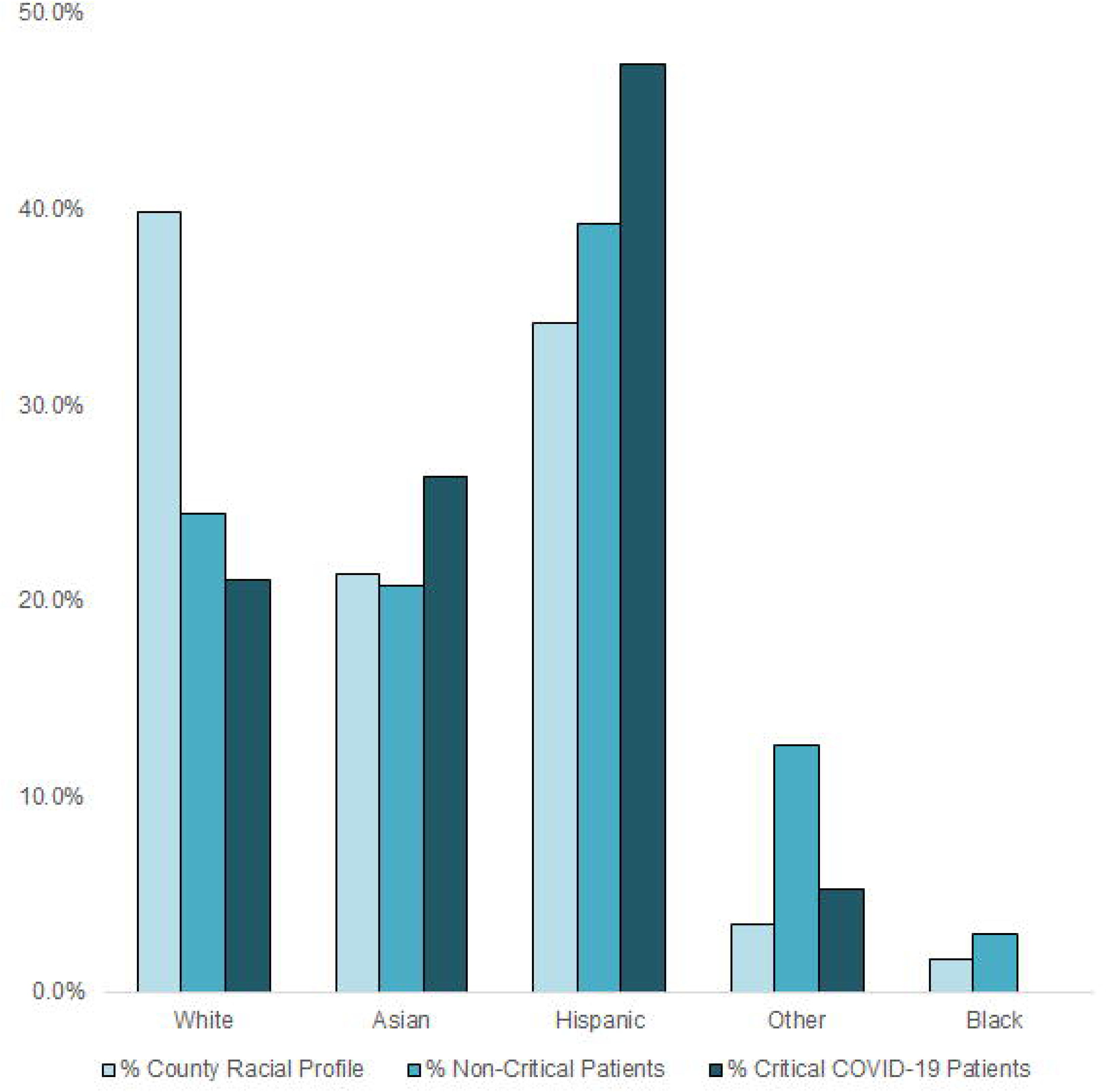
Race-ethnic disparities of COVID-19 patients. Bar charts demonstrating disparities between the Orange County racial profile and the distribution of Non-Critical COVID patients as well as Critical COVID patients at our institution. Specifically, while Hispanic/Latinx patients account for 34.2% of the county population, they represent 47.4% of our institution’s critical cases. Conversely, while White patients account for 40.1% of the county population, they represent 21.1% of our institution’s critical cases.

## Discussion

While recent epidemiologic reports on COVID-19 have observed disparities in incidence, morbidity, and mortality among underserved and minority populations, including the Hispanic/Latinx community, there remains a poor understanding about which social determinants may contribute to these disparities. Included in this descriptive case series is information about the impact of individual and neighborhood-level determinants on COVID-19 incidence over time and severity. Our observation of exponential growth in Hispanic/Latinx COVID-19 cases who live in dense households and low-income communities highlights the importance of understanding community-level factors during a pandemic.

The extensive research on the relationship between social determinants and race-ethnic disparities in noncommunicable diseases, including heart disease and stroke, has largely been characterized by long-term exposures leading to differences in disease ourcomes^16,17^. However, the social determinants of infectious disease are less well-characterized. Our finding that COVID-19 cases in Orange County increased linearly in neighborhoods with higher housing density, lower educational attainment, and lower health coverage further underscore the importance of understanding contextual factors surrounding infectious disease outbreaks.

It has been suggested that individuals living in disadvantaged communities may be at greater risk of infectious diseases^10^. For example, a significant association between severity and lower educational attainment (individual and community) was observed in a study examining factors associated with poorer outcomes during the H1N1 pandemic of 2009^18^. Other studies suggest that neighborhoods defined as disadvantaged were associated with higher influenza-related deaths, hospitalizations, and poorer vaccination coverage^9-11,19^. Similarly, for COVID-19 the interaction of social determinants at multiple levels may lead to a “perfect storm” that disproportionately affects underserved populations by negatively impacting both rates of infection and outcomes.

This study generates several hypotheses to explain the impact of social determinants of health on the observed disparities in COVID-19 incidence and severity over time. We hypothesize that underserved populations living in lower-income census tracts are experiencing a spike in COVID-19 transmission due to increased housing density, employment in “essential/frontline” Job sectors, lower educational attainment, and limited health care access. For health care workers, this trend has been seen across the U.S. and has been attributed to their “frontline” status with exposure to COVID-19 patients^20^. We hypothesize that the lower income and minority populations that demonstrate a similar increase over time may also be experiencing this “frontline” spike due to barriers to social distancing.”

Social policies at the local, county, state, and national level, including universal face masking, self and family case isolation, social distancing, school cancellation, and voluntary or mandated shelter in place, may underlie the connection between social determinants and COVID-19 disparities.^21^ For example, while social distancing has been demonstrated to be a strong and effective preventive measure against COVID-19 transmission rates^22,23^, our data suggest that those living in communities where work at home is infrequent had higher COVID-19 case growth. Adherence to social policies is likely difficult for populations that do not have and cannot afford remote work options. For example, approximately 75% of New York frontline and essential workers, including healthcare, grocery & convenience stores, public transit, cleaning services, and delivery operations, are people of color^24^. We suggest that social policies incorporate a framework around social determinants that would promote strategies to achieve equity for all communities.

In addition, pre-existing disparities in co-morbidities associated with disproportionately worse COVID-19 outcomes may be partially responsible for the race-ethnic disparities observed in this study. Data from the National Health and Nutrition Examination Survey (NHANES) suggests that more than a third of all US adults met the definition of metabolic syndrome with greater prevalence among Black and Latinx populations and in low socioeconomic populations^25^. The significantly greater presence of metabolic syndrome in underserved populations^11^ may provide some insight into the racial-ethnic disparities in COVID-19 incidence, particularly for critical disease

Limitations of this study include the small sample of patients from a single-center, so conclusions may not be broadly generalizable. However, the strengths of this study include the collection of comprehensive data, including race-ethnic and census-tract derived community determinants from all patients presenting with COVID-19 over the study period. Future studies should evaluate the complex interactions of the social determinants of income and ethnicity with other demographic, clinical, and laboratory factors.

In summary, our study examines the unveiling of race-ethnic disparities over the first six weeks of COVID-19 in Orange County, CA, and highlights vulnerable populations that are at increased risk for contracting COVID-19 and experiencing disproportionately severe outcomes. While our findings that Hispanic/Latinx populations are at increased risk corroborates reports elsewhere in the United States^2^, this study demonstrates the increase was most dramatic in minority groups living in disadvantaged communities. When we think of race-ethnic disparities, we often investigate immediate causes of disease, including risk factors. Our descriptive case series illustrates that for COVID-19 disparities, we also need to consider the “causes of those causes,” which ultimately set the stage for biological and social disparities.

## Data Availability

Data is recorded from the University of California, Irvine and available to researchers pending appropriate IRB requests.

